# Accounting for small-study effects using a bivariate trim and fill meta-analysis procedure

**DOI:** 10.1101/2020.07.27.20161562

**Authors:** Chongliang Luo, Arielle Marks-Anglin, Rui Duan, Lifeng Lin, Chuan Hong, Haitao Chu, Yong Chen

**Author notes:** Correspondence to: Yong Chen, Department of Biostatistics, Epidemiology and Informatics, University of Pennsylvania, Philadelphia, Pennsylvania 19104, USA.

## Abstract

In meta-analyses, small-study effects (SSE) refer to the phenomenon that smaller studies show different, often larger, treatment effects than larger studies, which may lead to incorrect, commonly optimistic estimates of treatment effects. Visualization tools such as funnel plots have been widely used to investigate the SSE in univariate meta-analyses. The trim and fill procedure is a non-parametric method to identify and adjust for SSE and is widely used in practice due to its simplicity. However, most visualization tools and SSE bias correction methods have been focusing on univariate outcomes. For a meta-analysis with multiple outcomes, the estimated number of trimmed studies by trim and fill for different outcomes may be different, leading to inconsistent conclusions. In this paper, we propose a bivariate trim and fill procedure to account for SSE in a bivariate meta-analysis. Based on a recently developed visualization tool of bivariate meta-analysis, known as the galaxy plot, we develop a sensible data-driven imputation algorithm for SSE bias correction. The method relies on the symmetry of the galaxy plot and assumes that some studies are suppressed based on a linear combination of outcomes. The studies are projected along a particular direction and the univariate trim and fill method is used to estimate the number of trimmed studies. Compared to the univariate method, the proposed method yields consistent conclusion about SSE and trimmed studies. The proposed approach is validated using simulated data and is applied to a meta-analysis of efficacy and safety of antidepressant drugs.

## 1. Introduction

Meta-analysis is a type of systematic review approach that uses statistical methods to synthesize evidence from multiple studies. In biomedical research, meta-analyses have been valued as the highest level in the hierarchy of evidence [4,22]. To conduct a meta-analysis, researchers need to comprehensively search for eligible studies in various databases, such as PubMed and the Cochran Library [5], to gather as complete a body of evidence as possible. It is often the case that smaller studies show different, often larger, treatment effects than larger ones, a phenomenon known as small-study effects. Common reasons for small-study effects include publication bias, outcome reporting bias and clinical heterogeneity. Small-study effects can negatively impact the validity of meta-analyses, as ignoring small-study effects and combining only the identified published studies or outcomes may lead to an incorrect, commonly optimistic conclusion.

Visualization tools have been developed to detect small-study effects in meta-analyses. In a univariate meta-analysis, a funnel plot is typically used to visualize the estimates from all studies, with the horizontal axis representing the estimated effect size and the vertical axis representing the precision (reciprocal of standard error) [1,24,25]. For bivariate outcomes, such as efficacy and safety measures in clinical trials, or the sensitivity and specificity in diagnostic tests, Hong et al. [3] proposed a new visualization tool, namely the galaxy plot, which can be viewed as an analog of the funnel plot in the bivariate case. Let *y*_*i*_ = (*y*_1*i*_,*y*_*2i*_)′ and (*S*_*i*_ = *S*_1*i*_,*S*_2*i*_)′ be the estimated effect sizes and their standard errors of the *N* studies in a bivariate meta-analysis (BMA) (*i* =1, *N*). In agalaxy plot, the *i* th study is represented by an ellipse, with the center at(*y*_1*i*_,*y*_2*i*_)′ the horizontal and vertical axes are proportional to the precision of two outcomes 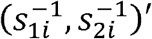. Figure 1 shows the funnel plot for a univariate meta-analysis and the galaxy plot for a BMA.

**Figure 1.**
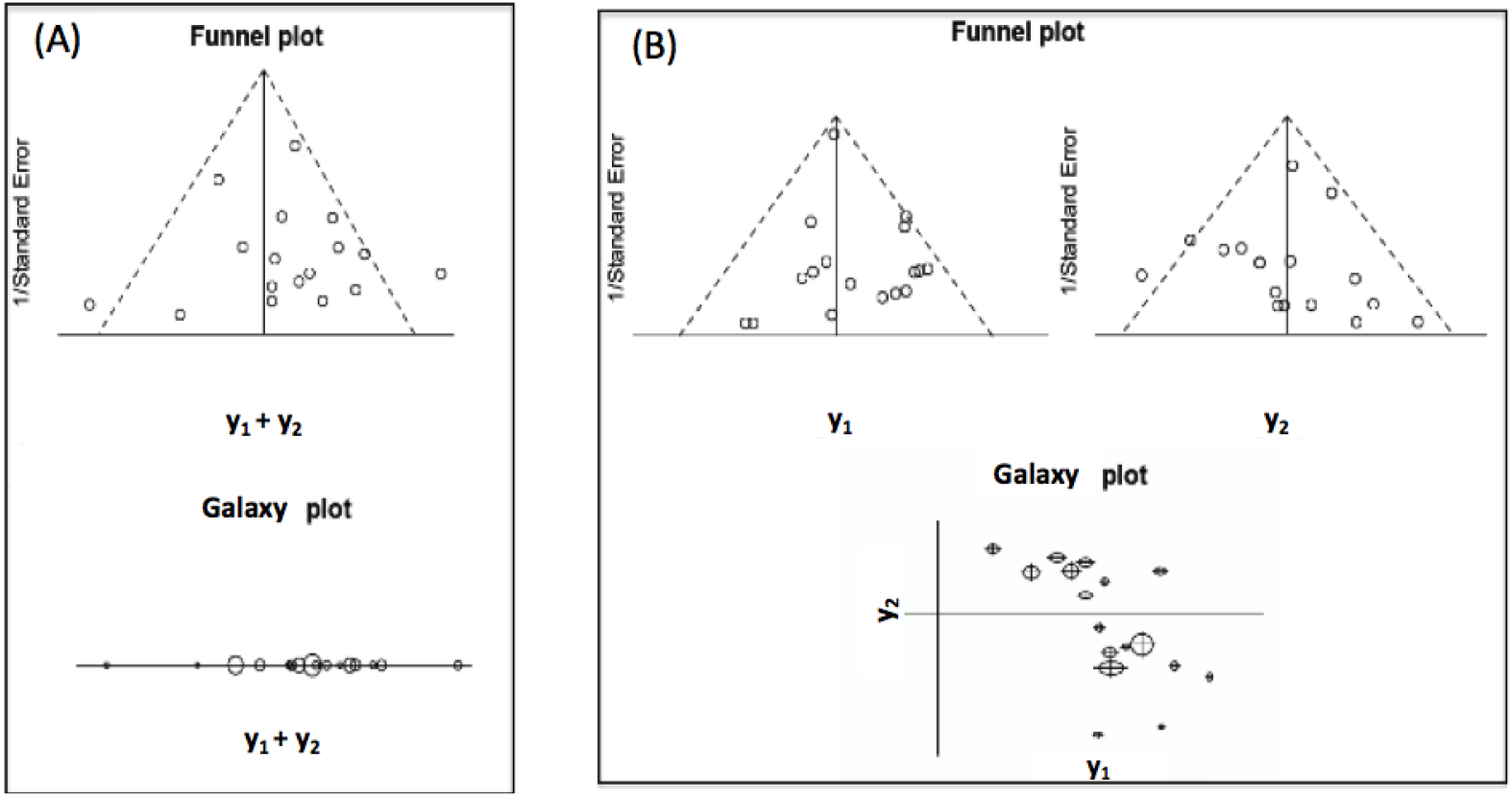
(A) The upper panel is the funnel plot for a univariate meta-analysis. The lower panel shows the galaxy plot reduced to a one-dimensional space, which retains all information of the funnel plot as the standard error is represented by each study’s circle radius. (B) The upper panel shows two separate funnel plots for the two outcomes in a bivariate meta-analysis. They can b integrated into the galaxy plot in a two-dimensional space in the lower panel.

Many statistical methods [5-9] based on the funnel plot or other visualization tools [13] have been developed to detect small-study effects. Among them, the trim and fill (T&F) method [9,17] is an attractive approach since it is nonparametric and only relies on the assumption of symmetry of the funnel plot. It imputes the potential unpublished studies and provides a bias-corrected estimate. It is worth mentioning that asymmetry of the funnel plot does not necessarily mean SSE, which could also be caused by the intrinsic correlation between the effect size and its standard error. However, after proper transformation, we can eliminate the intrinsic correlation and identify SSE by detecting asymmetry of the funnel plot [12]. Despite its popularity, the T&F method cannot be directly applied to bivariate meta-analysis. One naive solution is to apply the T&F method to each outcome separately, or to a weighted measure of the two outcomes. However, inconsistent conclusions may be drawn, as different studies may be trimmed when different marginal measures are considered.

Multivariate meta-analysis has recently received more attention [21], but few statistical methods have been developed to account for SSE in multivariate scenarios. Recently, Hong et al. [18] proposed a multivariate extension of Egger’s test for detection of SSE in multivariate settings. Compared to univariate Egger’s tests, the multivariate test yields a consistent conclusion of potential SSE and has superior power for identifying SSE by combining signals of SSE from multiple outcomes. Along this line of research, this paper proposes a SSE correction method based on the galaxy plot for bivariate meta-analysis. It is an extension of the univariate T&F method. It assumes symmetry of the galaxy plot in that studies on one side of a suppressing line are unpublished. The key idea is to project the bivariate outcome to one of a sequence of directions, and to use the univariate T&F method in estimating the number of suppressed studies. We choose the direction along which most studies are trimmed as the optimal projection direction. This is motivated by the intuition that compared to other direction, the most studies are expected to be trimmed along the true suppression direction. The identified suppressed studies are then filled (i.e. imputed) in the galaxy plot by symmetry, and the final effect size is estimated from the observed and the filled studies.

To the best of our knowledge, the proposed bivariate T&F method is the first attempt to use imputation-based nonparametric methods for bias correction in multivariate meta-analysis. It relies on the symmetry of the galaxy plot and yields a consistent conclusion about SSE, identifying possible linear combinations of outcomes that the missing studies are based on. The method is a useful approach for sensitivity analysis of SSE. The rest of this paper is organized as follows. Section 2 provides the formal description of the proposed method, and Section 3 demonstrates the performance of the method using simulated data. We apply the method to a real-world meta-analysis of efficacy and safety of antidepressant drugs in Section 4. We discuss limitations in Section 5.

## 2. Methods

Assume studies are suppressed based on a weighted score of the two outcomes, i.e. a linear combination *z*_*i*_ =*c*_1_*y*_1*i*_+*c*_2_*y*_2*i*_,*i* = 1,…, *n*. The studies with the smallest weighted scores are suppressed The values of *c*_1_ and *c*_2_ or essentially the ratio of *c*_1_ and *c*_2_ can be pre-specified depending on the outcomes or can be decided from the data. For example, in a meta-analysis of diagnostic tests, the two outcomes are the sensitivity and the specificity, thus *z*_*i*_ is equivalent to the Youden’s index [19] when *c*_1_ and *c*_2_. For severe diseases such as cancers, *c*_1_ > *c*_2_ can be chosen to reflect the higher penalty on false negative than false positive conclusions.

When it is infeasible to pre-specify the values, we propose a searching algorithm to find the optimal ratio of *c*_1_ and *c*_2_, which gives the most trimmed studies. This is based on the observation that the closer a direction is to the truth, the more studies are expected to be trimmed along that direction. We set a sequence of angles 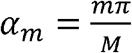 The step-by-step bivariate T&F procedure is described as follows:

1. For each 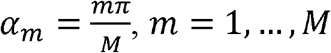, M is the number of directions, and ρis the correlation between two outcomes, both are pre-specified.
  a. Calculate the weighted scores and variances, 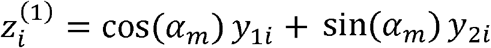 and 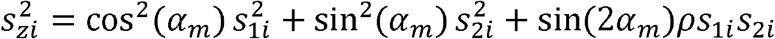 for each study *i*, and the center 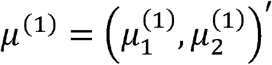 and its associated score 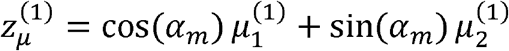
  b. Based on 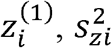, and 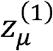, estimate the number of suppressed studies as 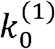
  c. By the symmetry assumption, trim 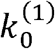 studies with the largest values of 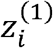
  d. Update the center and its weighted score as *μ*^(2)^and 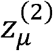, using the remaining studies.
  e. Repeat steps b)–d) until iteration *J* such that no more studies are trimmed, i.e.,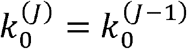
  f. Fill the galaxy plot by point symmetry about the center *μ*^(*J*)^, i.e., 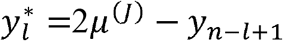, with equal standard errors 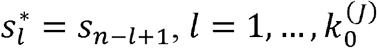
2. Denote 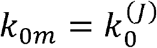 from step 1), and estimate the center as *μ*_*m*_ based on the observed and filled *n* + *k*_0m_ studies.
3. Select *m*\* that gives the largest *k*_0m_; the bias-corrected center is *μ* _m_ *

The algorithm is also demonstrated by Figure 2, using simulated studies. We have the following remarks on the proposed procedure. First, in (a), deriving the variance of the weighted score requires a pre-specified correlation. *ρ* We can use the estimated correlation between the two outcomes 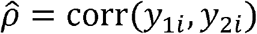 as a proper value. Further sensitivity analysis can be performed to evaluate the impact of different values of. In (b), we can use one of the estimators in the univariate T&Fmethod, i.e. *R*_0_, *L*_0_, or *Q*_0_ to estimate the number of suppressed studies, as described by Duval and Tweedie [9, 17]. As the result of the T&F method depends on the estimators being used, it is suggested to present the three estimators as a sensitivity analysis [1,2,9,14]. Meanwhile in (d), either a random-effect or fixed-effect model can be used to estimate the center. However, when both the publication bias and between-study heterogeneity exist, the random-effect model tends to give larger weights to the smaller studies and thus induce larger bias for the effect size estimation [2]. We thus always use a fixed-effects model for updating the center in (d), and use a random-effects model for the filled studies in (2). This is referred to as FE-RE T&F method [2,15]. Finally in (3), there may be ties for the values of {*k*_0*m*_, *m* = 1, …, *M*}. If the values of for ties are clustered together, we can choose the median of the cluster as the optimal angle. For example, if *m* = 1, 2, 3, 5 all produce the same maximum value of *k*_0*m*_, we may choose *m*\* = 2. Otherwise, we can also increase *M* to find a better decision for the ties. On the other hand, the ties can help us evaluate the assumption of linear suppression. If many ties continue to exist with larger *M*, they imply that the studies are less likely to be suppressed along a specific direction.

**Figure 2.**
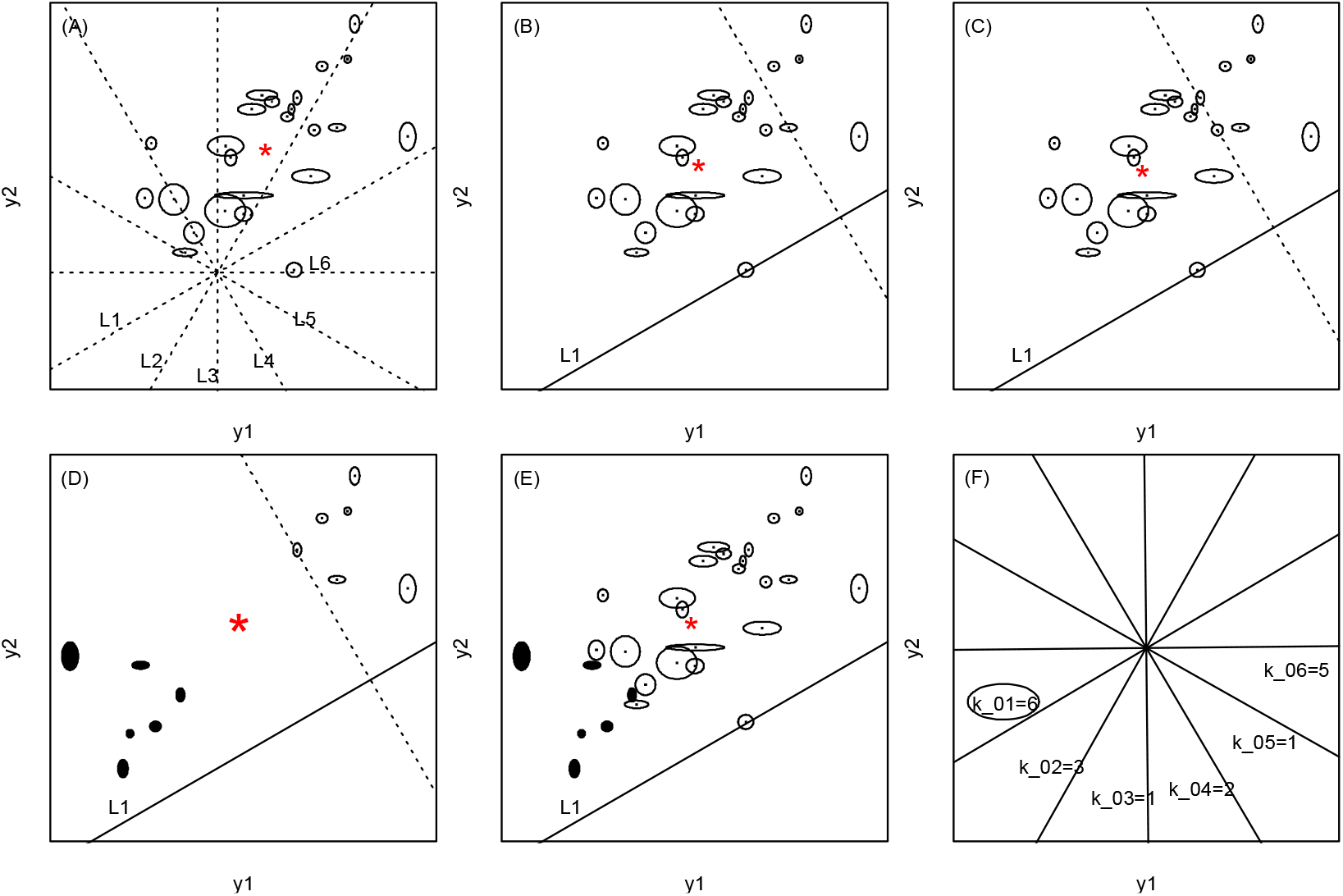
Bivariate trim and fill method to identify and correct for small-study effects in the galaxy plot of a bivariate meta-analysis. (A) The galaxy plot shows 24 simulated studies and the dashed lines represent M=6 directions. The red star is the center. (B) For the first direction L_1_, the first iteration trims 4 studies with the largest weighted scores, using the univariate trim and fill method. The red star represents the updated center using the 22 studies. (C) The second iteration trims two more studies and the red star represents the updated center using the 20 studies. No more studies are trimmed in the next iteration so the number of trimmed studies is *k*_01_ = 6. (D) 6 studies are filled by point symmetry about the center, with equal standard errors. (E) The center is estimated based on the 30 fully augmented studies. (F) Repeated steps as in (B)–(E) for other directions with {*k*_0*m*_, *m* = 2,…, *M*}, implying that *L*_1_ is the optimal direction since *k*_01_ is the largest.

## 3. Simulation study

We demonstrate the proposed approach using simulated bivariate meta-analyses. The number of observed studies is *n* = 25,50,100 and the number of Suppressed studies is.*k*_0_ = 10,20 We generate the bivariate outcome and its variance according to a random-effect model:

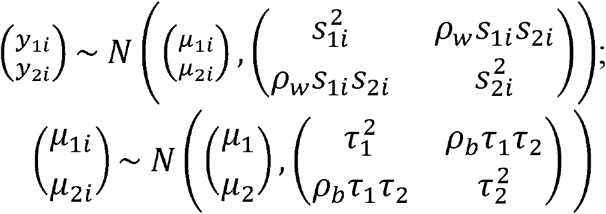

for *i*=1,…,*n*+*k*_0_ where 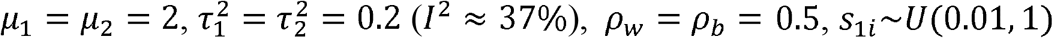 and *s*_2*i*_ ∼*U* (0.01,1) The *k*_0_ studies with the smallest value of 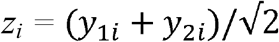 are then suppressed. We conduct the bivariate T&F along each of the M=12 projection directions and compare the adjusted effect size estimation along the selected direction (tf.biv) with unadjusted multivariate meta estimation (unadjusted) and the univariate T&F (tf.uni) estimation. To evaluate the searching algorithm’s performance, we also compare the proposed approach with the bivariate T&F along the true suppression direction (tf.biv.true). We only present the result of T&F using the *R* _0_ estimator, as the *L* _0_ and *Q*_0_ estimators give similar results in this simulation study.

The results of a typical setting are presented in Figure 3. The galaxy plot in (A) shows a typical simulated dataset, with 50 studies being observed and 20 suppressed. In (B), the proposed bivariate T&F method leads to better correction towards the true effect size, compared to the univariate T&F. This is because the univariate T&F didn’t fully utilize the symmetry of the galaxy plot. Notice that the bivariate T&F with the proposed searching algorithm can obtain similar amount of bias reduction compared to the bivariate T&F along the true direction. This is more explicitly explained in (C) and (D). In (C) the T&F along the true direction tends to trim more studies compared to other directions and as a consequence, in (D) more than 50% of the time the searching algorithm selected the true direction as optimal. The results of bias correction and number of trimmed studies with various number of simulated studies are presented in Table 1. Since the bias correction for the two outcomes are similar, only the results of the first outcome are shown.

**Table 1.** Simulation study results. The number of observed and suppressed studies are n and k_0_ respectively. Listed are the bias of the estimation of the first outcome’s effect size 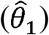, and number of suppressed studies 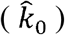 for unadjusted multivariate meta-analysis (mvmeta), univariate T&F (tf.uni), proposed bivariate T&F along the true projection direction (tf.biv.true) and the proposed bivariate T&F with direction-searching algorithm (tf.biv). The Monte Carlo standard errors are also listed in the parentheses.

| n | $k_0$ | unadjusted | tf.uni | | tf.biv.true | | tf.biv | |
| --- | --- | --- | --- | --- | --- | --- | --- | --- |
| | | $\hat{\mu}_1$ (s.e.) | $\hat{\mu}_1$ (s.e.) | $\hat{k}_0$ (s.e.) | $\hat{\mu}_1$ (s.e.) | $\hat{k}_0$ (s.e.) | $\hat{\mu}_1$ (s.e.) | $\hat{k}_0$ (s.e.) |
| 25 | 10 | 0.27 (0.15) | 0.17 (0.24) | 4.55 (3.53) | 0.09 (0.22) | 6.16 (5.57) | 0.07 (0.27) | 9.27 (6.23) |
| 25 | 20 | 0.41 (0.15) | 0.30 (0.23) | 5.05 (3.79) | 0.25 (0.20) | 7.26 (5.46) | 0.22 (0.28) | 10.67 (6.23) |
| 50 | 10 | 0.16 (0.10) | 0.07 (0.20) | 7.53 (6.00) | 0.01 (0.17) | 9.35 (8.31) | -0.02 (0.18) | 11.87 (8.82) |
| 50 | 20 | 0.27 (0.10) | 0.16 (0.18) | 8.25 (6.22) | 0.07 (0.18) | 14.55 (10.71) | 0.06 (0.21) | 17.27 (11.6) |
| 100 | 10 | 0.09 (0.07) | 0.03 (0.16) | 10.54 (8.81) | -0.01 (0.10) | 10.63 (8.01) | -0.02 (0.13) | 12.17 (8.85) |
| 100 | 20 | 0.17 (0.07) | 0.08 (0.15) | 12.12 (9.90) | 0.00 (0.13) | 20.78 (14.39) | -0.01 (0.14) | 21.8 (14.44) |

**Figure 3.**
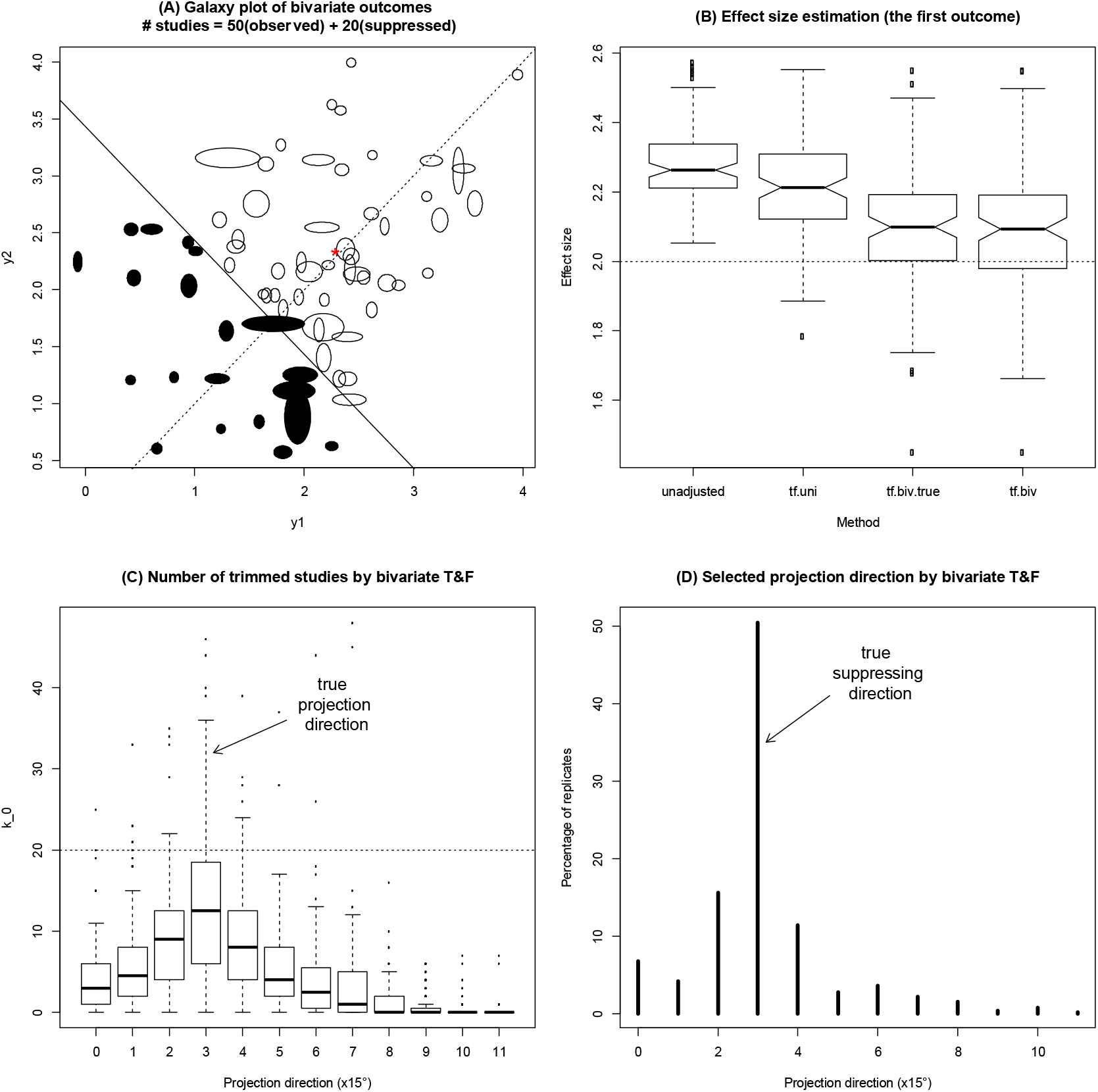
Simulation study results of 200 replicates of meta-analyses, each containing 50 observed studies after suppressed 20 studies. (A) The galaxy plot showing a typical simulated bivariate meta-analysis dataset according to a random-effect model. The 70 studies are projected to the true projection line (dashed line, 45° to the x axis) among which 20 studies with the smallest projected values (to the lower left of the solid line) are suppressed and the other 50 studies (to the upper right of the solid line) are observed. (B) The effect size estimation of the unadjusted multivariate meta-analysis (unadjusted), the univariate T&F (tf.uni), the bivariate T&F along the true projection direction (tf.biv.true) and the bivariate T&F using the searching algorithm (tf.biv). (C) The estimation of the number of suppressed studies (k_0_) along each of the candidate projection directions. The true value is 20. (D) The selected projection direction by the searching algorithm. Lines represent the percentages of times each direction was selected (with the most studies been trimmed) among the 200 replicates.

## 4. Real data application

We apply the proposed bivariate T&F approach to a recently published meta-analysis of randomized controlled trials of 21 antidepressants [16]. We focus on the direct comparison between antidepressants and placebo, and omit the drug-drug comparison. The comparisons of any the antidepressant drug versus placebo are pooled together to form a meta-analysis of “active treatment” with antidepressants. Similar pooling approach has been used in Chaimani et al. [23] when there are multiple treatments. The analysis includes published studies as well as some “grey literature” studies that can be used to validate the potential publication bias. The total number of studies is 377, among which 289 are published and 88 are collected from grey literature. The two outcomes are efficacy (whether response to treatment) and safety (whether dropout due to any reason). Since the original outcomes are binary, we adopt the arcsine transformation [12] to remove the intrinsic correlation between study-specific effect size estimates and their sample variances. The final outcomes are the arcsine difference of response rates of drugs vs placebo, and the arcsine difference of dropout rates of placebo vs drugs. The greater the two outcomes are, the better the treatment is.

As in the simulation study, we use FE-RE T&F approaches and present the results of R_0_ and L_0_ estimators. Figure 4 shows the number of trimmed studies along the 12 directions of projection and correspondingly Table 2 shows the estimated effects. As seen from Table 2, the numbers of trimmed studies are not consistent by using univariate T&F on the two outcomes, while the proposed bivariate T&F gives one unique number of trimmed studies along a specific direction. In Figure 4, the galaxy plot of the published studies is in the left panel, and the galaxy plot of both published and grey literature studies is in the right panel. Generally speaking, studies are more likely to be missing if either or both of the two outcomes are small, i.e. at the bottom left corner. This can be seen by the estimates from Table 2 and also the trimmed studies in Figure 4.

**Table 2.**
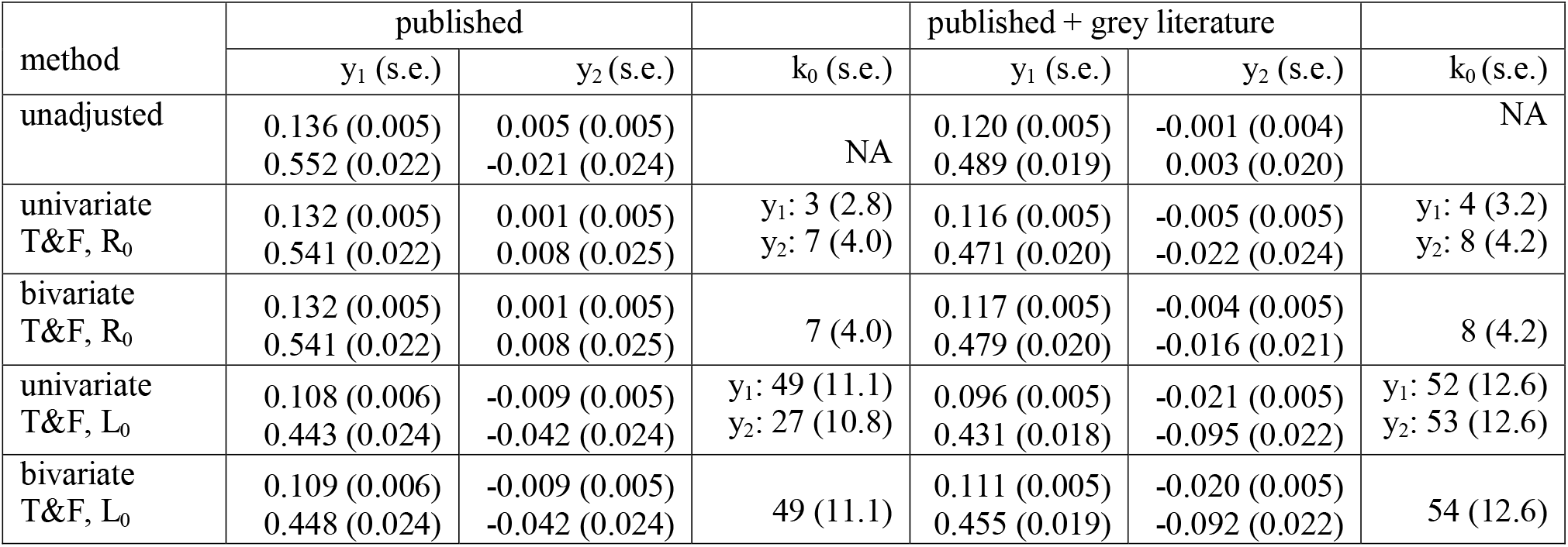
The estimated effects of efficacy (y_1_) and safety (y_2_) outcomes in the meta-analysis of antidepressant drugs using unadjusted, univariate and proposed bivariate T&F approaches, with standard errors in the parentheses. Larger effects favor treatment over control. In each cell the upper is the outcome on the arcsine-difference scale and the lower is the outcome on the (log) OR scale. The T&F method is conducted on the arcsine-difference scale and the (log) OR is then calculated based on the filled studies.

**Figure 4.**
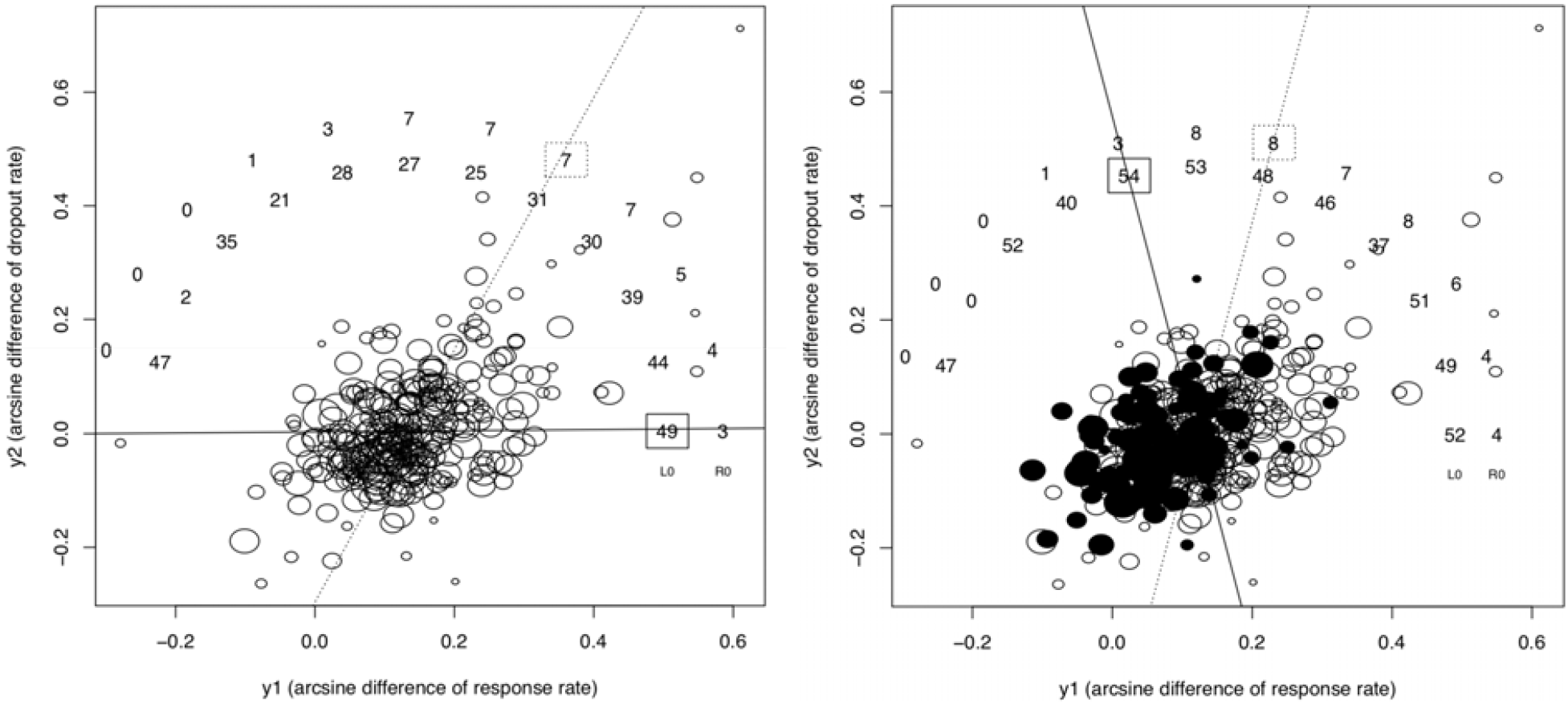
The number of trimmed studies by the proposed bivariate T&F method in the antidepressant meta-analysis of Cipriani et al [16]. The two outcomes are the arcsine difference of response rate and dropout rate between antidepressants and placebo. The 288 published studies are presented in the galaxy plot in the left panel while 89 “grey literature” studies (colored) are additionally included in the right panel. The number of trimmed studies along the 12 directions of projection are shown, where the inner and outer circles are for L_0_ and R_0_ estimators respectively. The direction with maximum number of trimmed studies is selected.

In the left panel of Figure 4, the bivariate T&F using L_0_ estimator trims 49 studies on the right along the horizontal direction, which is more than other directions. This selected direction indicates that the studies are missing primarily due to insufficient evidence of efficacy. Meanwhile, using R_0_ estimator, only 7 studies are trimmed on the top-right corner along several directions with tied number of trimmed studies. The discrepancy between the two estimators is because the missing mechanism assumption of T&F is not strictly satisfied in this real-world example and thus the R_0_ estimator can only trim those studies with extreme outcome values [9]. Nevertheless, T&F using the R_0_ estimator shows that some studies are missing with insufficient evidence of both efficacy and safety. On the other hand, in the right panel where both the published and grey literature studies are used, T&F using the L_0_ estimator selects a different direction than that in the left panel. However, the variation of the number of trimmed studies is also much smaller than that in the left panel. This shows that the “grey literature” has adjusted much of the bias due to missing studies and the remaining bias is not along a specific direction. The bias correction for both outcomes is also shown in Table 2, where the unadjusted estimate of efficacy outcome using published and grey literature studies is close to the adjusted estimate, by the bivariate T&F with L_0_ estimator using either published or published plus grey literature studies. Lastly, notice that in Figure 4, T&F with the R_0_ estimator using published and grey literature studies obtains a similar number of trimmed studies along a similar direction as that of using published studies only. This indicates that the grey literature studies don’t adjust for the bias due to the missing studies with extreme values of both outcomes.

## 5. Discussion

We proposed a trim and fill method for bivariate meta-analysis to correct for potential small study effects. This bivariate T&F method is based on the galaxy plot, which is an analog of the funnel plot for bivariate meta-analysis. By trimming and filling the studies that are projected onto a sequence of directions, the method can detect potential directions where studies are most likely to be unpublished or missing. When the missingness of studies depends on more than one outcome, the method can achieve a potentially better bias reduction than the univariate T&F method. The method’s performance is demonstrated using a simulation study and a real-world meta-analysis of efficacy and safety of antidepressant drugs. This large-scale meta-analysis example shows that studies are missing primarily due to efficacy, while some missing studies are possibly due to both efficacy and safety. After including the studies collected from grey literature, much of the bias is corrected. However, the bias due to missing studies with both extreme response and dropout rates are not likely to be corrected. The corrected effect size estimates using either published or published and grey literature studies are consistent.

Some well-known limitations of the univariate T&F method still remain in the bivariate scenario. For example, the method is sensitive to outlying studies. As a result, it is recommended that both L_0_ and R_0_ estimators should be used [9,14,17]. In the real data example, it is obvious that the missing studies are not suppressed by a straight line, thus using L_0_ gets more trimmed studies than using R_0_. A more essential limitation is that, when between-study heterogeneity is severe, the T&F method usually fails to identify missing studies and correct for the bias [2,20]. In our simulation and real data examples, the heterogeneity is not severe (*I*^2^ ≈ 37% in simulation and 38% in real data). The FE-RE T&F approach, as we adopted in this paper, can alleviate this problem. While the proposed bivariate T&F method is a useful tool for sensitivity analysis of small-study effects, it mainly works for publication bias and cannot account for outcome reporting bias. This could be an interesting future research direction.

- What is already known Trim and Fill method is one of the methods to account for small study effects in meta-analysis, despite several known limitations, such as sensitive to moderate or large between-study heterogeneity.
- What is new Trim and Fill method is not available for bivariate or multivariate meta-analysis where multiple outcomes are jointly considered. In this paper, we propose a bivariate trim and fill procedure to account for small study effects in bivariate meta-analysis. This procedure is based on a recently proposed visualization tool for bivariate meta-analysis data, known as galaxy plot, and jointly imputes the missing studies based on the point symmetry of the studies in bivariate visualization.
- Potential impact for RSM readers outside the authors’ field

The proposed method is nonparametric (i.e., makes minimal model assumptions) and easy to understand for non-technical researchers who are interested in conducting sensitivity analyses or bias corrections for small study effects in bivariate meta-analyses.

## Data Availability

The antidepressant drug data is published and available from the Lancet.

https://www.thelancet.com/journals/lancet/article/PIIS0140-6736(17)32802-7/fulltext

## Notes

### Competing Interest Statement

The authors have declared no competing interest.

### Funding Statement

NO funding related.

